# Epidemiology, determinants, and microbiological profile of implant-associated infections following intramedullary nailing in a tertiary hospital in Ethiopia

**DOI:** 10.1101/2025.09.22.25336390

**Authors:** Abebaw Muhabaw Zegeye, Ephrem Bekele Kebede, Birlew T Alemayehu, Sileshi Serebe Zeleke, Kemal Tesfa Alemu, Yohannis Derbew Molla

**Affiliations:** Department of Orthopedics, University of Gondar, College of Medicine and Health Sciences, Gondar, Ethiopia; Department of Internal Medicine, University of Gondar, College of Medicine and Health Sciences, Gondar Ethiopia; Department of General Surgery, University of Gondar, College of Medicine and Health Sciences, Gondar, Ethiopia

**Keywords:** intramedullary nailing, implant-associated infection, long bone fractures, epidemiology, determinants

## Abstract

**Background:** Implant-associated infections after intramedullary nailing (IMN) are a major cause of morbidity in patients with long bone fractures. This study aimed to determine the magnitude, determinants, and microbiological profile of such infections in a tertiary hospital in Ethiopia.

**Methods:** A hospital-based cross-sectional study was conducted among 260 patients who underwent IMN for lower extremity long bone fractures between January 2021 and December 2023. Clinical data, operative details, and microbiological results were collected, and logistic regression was applied to identify determinants of infection.

**Results:** The overall infection rate was (8.46% (95% CI: 5.38–12.53) (22/260), with 4.62% deep and 3.85% superficial infections. Emergency surgery and hospital stay >14 days were independently associated with increased infection risk. All culture isolates were Gram-negative bacteria, with Escherichia coli being the most frequently identified pathogen.

**Conclusion:** The infection rate was higher than in many other settings, underscoring the need for improved perioperative infection prevention and reduced hospital stays. Findings highlight the importance of perioperative optimization and hospital infection control in LMICs.

## Introduction

Lower extremity long bone fractures, particularly of the femur and tibia, are among the most common injuries encountered in orthopedic and trauma centers and represent a leading cause of hospital admission(1,2). These fractures also impose a substantial social and economic burden(3). They frequently result from high-energy mechanisms such as road traffic accidents, falls from height, and ballistic trauma(1,2,4,5).

Intramedullary nailing (IMN) is the preferred surgical technique for stabilizing these fractures because of its favorable biomechanical properties and relatively low complication rates(1,6–10). Nevertheless, implant-associated infections remain a serious postoperative complication, leading to increased morbidity, prolonged hospitalization, higher healthcare costs, and, in severe cases, revision surgery or limb loss(11–13).

Globally, studies on infection following IMN have generally focused on specific bones (femur or tibia) or fracture types (open versus closed). Reported infection rates vary widely. A multicenter prospective cohort study estimated an overall infection rate of 2.5% following tibial and femoral IMN surgeries(14). Similarly, retrospective cohort studies have reported rates of approximately 6.7% after IMN for long bone fractures(9). For open fractures, the risk is higher: a systematic review of 32 studies involving 3,060 patients reported infection rates of 5–16% for open tibial shaft fractures(15), while a meta-analysis of 17 studies on open femoral shaft fractures found rates of 5–6% (6). In general, studies of fracture fixation surgeries have reported infection rates between 5% and 6%(3,16).

In low– and middle-income countries (LMICs), a large multicenter study analyzing 46,113 IMN procedures reported relatively low crude infection rates of 0.8–1.5%, with femoral fractures less affected than tibial fractures(7). However, these figures likely underestimate the true burden due to methodological limitations such as incomplete follow-up and underreporting, as many patients in resource-constrained settings do not return unless complications arise. Indeed, when only patients with documented follow-up were included, infection rates increased substantially—up to 6.9% for tibial fractures—suggesting that the actual burden in LMICs may be higher than crude estimates indicate(7).

In Ethiopia, evidence remains limited. The few studies available have reported infection rates ranging from 4% to 9% following IMN for lower extremity long bone fractures(17,18). However, these studies were restricted in scope and did not explore in detail the risk factors or microbiological characteristics of implant-associated infections.

Understanding the microbiological profiles of pathogens responsible for implant-associated infections, as well as their antimicrobial susceptibility patterns, is critical for guiding empirical antibiotic therapy and informing infection prevention strategies. Furthermore, multiple risk factors—including high-grade open fractures (Gustilo-Anderson), delayed surgical intervention beyond 24 hours, tobacco use, diabetes mellitus, prolonged operative time, and omission of prophylactic antibiotics—have consistently been associated with an increased risk of infection(7,9,14,16). Yet, little is known about the relative importance of these factors in the Ethiopian context.

To address these gaps, this study was conducted to determine the magnitude of implant-associated infections following IMN surgeries for long bones at a tertiary hospital in Ethiopia, to identify patient, injury, and perioperative factors associated with infection, and to describe the microbiological profile and antimicrobial resistance patterns of pathogens isolated from infected cases. By generating such evidence, this research seeks to support clinical decision-making, optimize perioperative care, and inform infection prevention strategies in resource-limited settings.

## Methods and Materials

Study setting and Design: This hospital-based cross-sectional study was conducted at the University of Gondar Comprehensive Specialized Hospital (UOG CSH), a tertiary referral center located in Northwest Ethiopia.

Study Population: The study population included all patients who underwent IMN surgeries for fractures of the femur or tibia between January 2021 and December 2023.

Eligibility Criteria: Patients were eligible for inclusion if they received IMN surgery for tibial or femoral fractures at UOG CSH during the study period. Patients referred to UOG CSH after undergoing IMN surgery at other institutions, as well as those with pathological fractures, were excluded.

Sample size and Sampling Technique: A total of 295 patients who underwent IMN surgery were initially identified. Of these, 6 patients were excluded: 3 due to pathological fractures and 3 who were referred from other hospitals after undergoing IMN surgery. This yielded 289 eligible cases. Subsequently, 29 patients were excluded due to incomplete data registry, resulting in a final analytic sample of 260 patients (Figure 1).

**Figure 1:**
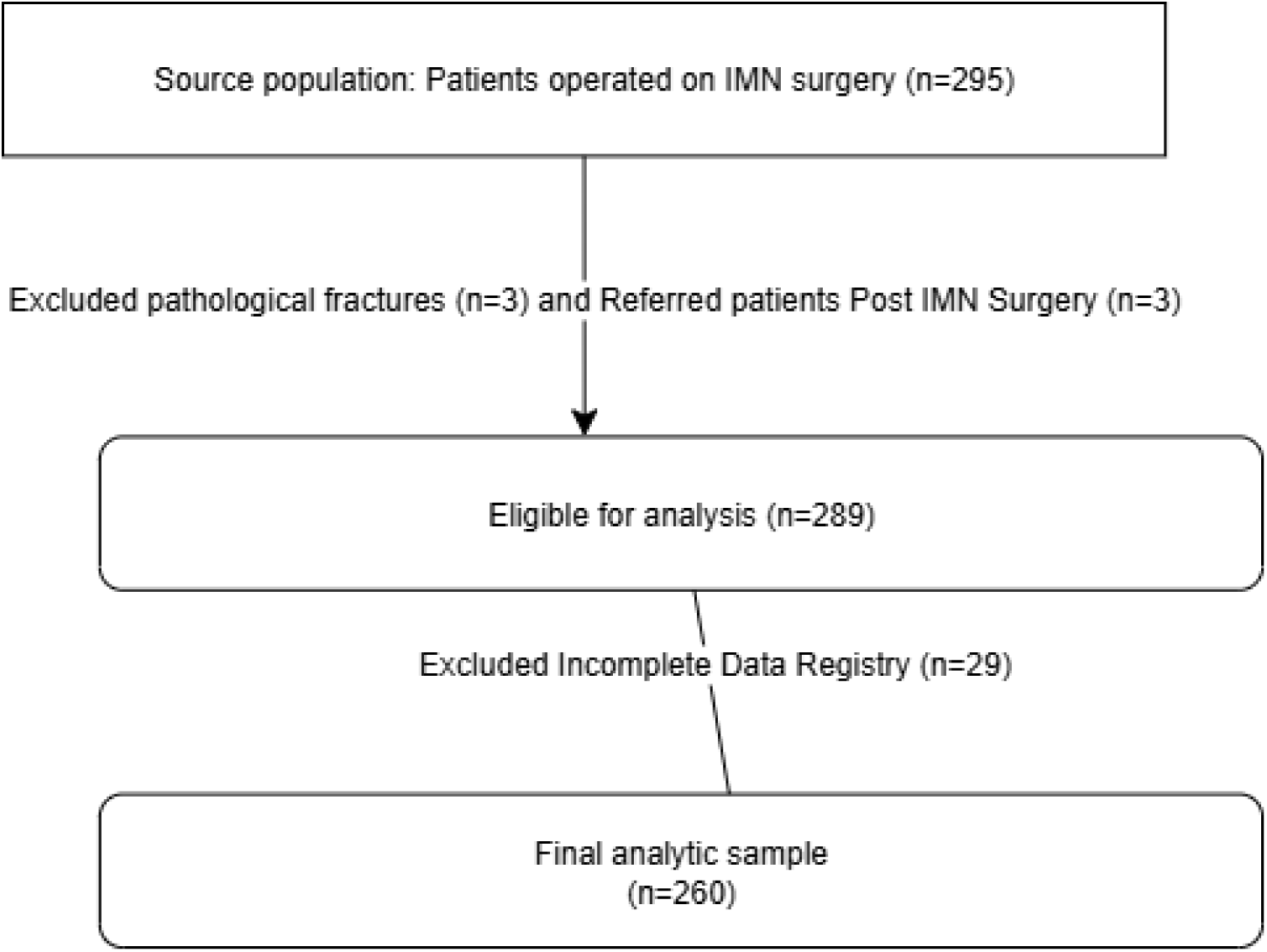
Flow diagram showing patient selection process.

Study Variables: The primary outcome variable was the occurrence of implant associated infection following IMN surgery, documented by the operating surgeon based on Metsemakers et al criteria(19).

Independent variables included sociodemographic characteristics (age, sex, residence, and occupation), clinical presentation factors (systemic comorbidity status, initial hemoglobin level, and time from injury to hospital presentation), injury-related characteristics (mechanism of injury, polytrauma status, bone fractured, fracture type, and Gustilo-Anderson classification for open fractures), and perioperative management variables (timing of antibiotic initiation and initial debridement for open fractures, staging of IMN surgery, timing of definitive IMN surgery, type of IMN surgery, reduction technique for closed fractures, and duration of hospital stay).

**Operational definitions**: **Intramedullary nailing** was defined as a major orthopedic procedure involving the insertion of a metallic rod into the medullary cavity of the femur or tibia, secured with proximal and distal interlocking screws.

**Implant-associated infection** was defined based on the criteria by Mestemaker et al(19). Accordingly, an infection was considered present if the operating surgeons documented clinical features such as fistula, sinus tract, or wound breakdown communicating with bone or implant; purulent drainage or intraoperative pus; or microbiological confirmation of identical pathogens from at least two separately collected deep tissue or implant specimens. Moreover, since this was a retrospective chart review, all cases labeled as implant-associated infection in the medical records by the operating surgeon were accepted under the assumption that the attending surgeons applied these criteria.

**Emergency IMN Surgery**: For this study, emergency surgery was defined operationally as any IMN procedure documented as “emergency” in the operation note, regardless of the underlying clinical indication. This included cases performed within a short timeframe after hospital presentation due to urgent clinical need (e.g., open fracture, neurovascular compromise, compartment syndrome) as well as procedures expedited for institutional or logistical reasons (e.g., day-time availability of the operating theater, limited elective slots).

**Elective IMN Surgery**: Elective surgery was defined as any IMN procedure documented as “elective” in the operation note, generally performed after initial stabilization and scheduling, with sufficient time for preoperative evaluation and preparation.

**Data Collection Procedures and Ethical Clearance**: Patient chart review was conducted from February 05, 2024, to May 30, 2024. Data was collected retrospectively from patient medical records using a pretested structured questionnaire. None of the authors had access to patient identifiers; all data were fully de-identified prior to analysis. Ethical clearance was obtained from the Institutional Review Board (IRB) of the UOG, and administrative permissions were secured prior to data collection.

**Data management and analysis**: Following data collection, entries were coded and input into EpiData version 3.1, then exported to Stata version 17.0 for statistical analysis. Data cleaning procedures were performed to ensure accuracy, consistency, and completeness. Descriptive statistics were presented using frequencies, proportions, and graphical summaries.

Binary logistic regression was employed to identify factors associated with implant-associated infections. Candidate Predictor variables for multivariable logistic regression were selected based on bivariate analysis (P≤0.2). Variables with sparse observations-such as Staged IMN surgeries, comorbidity status, smoking history, and closed reduction technique were excluded due to insufficient observation (< 5 % of total cases), which could compromise model stability. During multivariable logistic regression, certain categories such as bullet injury under the variable mechanism of injury were omitted due to perfect prediction, which prevented reliable estimation of adjusted odds ratios. These omissions were documented and interpreted accordingly in the Results section. Collinearity was assessed using variance inflation factors (VIFs) and linear regression diagnostics. The variable Closed vs Open fracture was excluded due to its collinearity with Gustilo-Anderson grade of the open fractures, which was retained for its broader clinical relevance and stronger statistical performance. The variables retained in the final multiple logistic regression model included: age group, duration of injury at presentation, GA grade of the open fractures, time to initial debridement, type of IMN surgery (elective vs emergency), duration of hospital stay, mechanism of injury and poly trauma status(Table 3).The strength of association between independent variables and the outcome was assessed using odds ratios (OR) with 95% confidence intervals (CI), and statistical significance was set at p < 0.05.

## Results

### Socio-Demographic and Clinical Characteristics

This study included a total of 260 patients who underwent intramedullary nailing (IMN) surgeries for tibial or femoral fractures between January 2021 and December 2023 at the UOG CSH. Age was categorized into three groups—young adults (18–30 years), middle-aged adults (31–60 years), and geriatric patients (≥61 years)—to facilitate subgroup comparisons and account for the non-normal distribution of age. Among these, nearly 60% were young adults aged 18–30 years, while only 17 patients (6.54%) belonged to the geriatric age group. A substantial proportion—196 patients (75.38%)—resided in rural areas. In terms of sex distribution, 217 patients (83.46%) were male, and 43 (16.54%) were female, reflecting the predominance of male trauma cases in the study setting. Regarding occupation, nearly half of the participants (124, or 47.69%) were farmers, followed by students, housewives, others and merchants (Table 1).

**Table 1:**
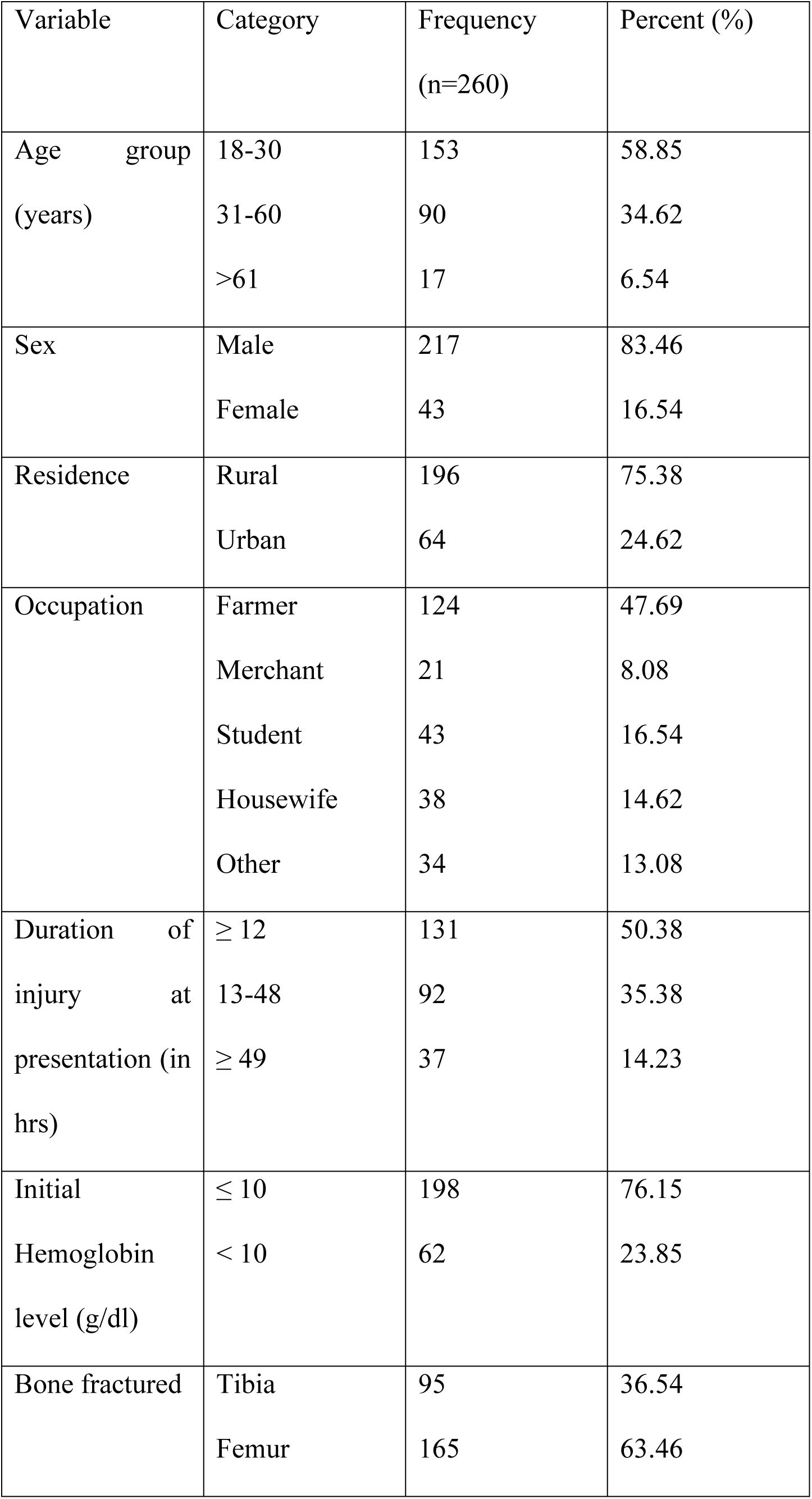
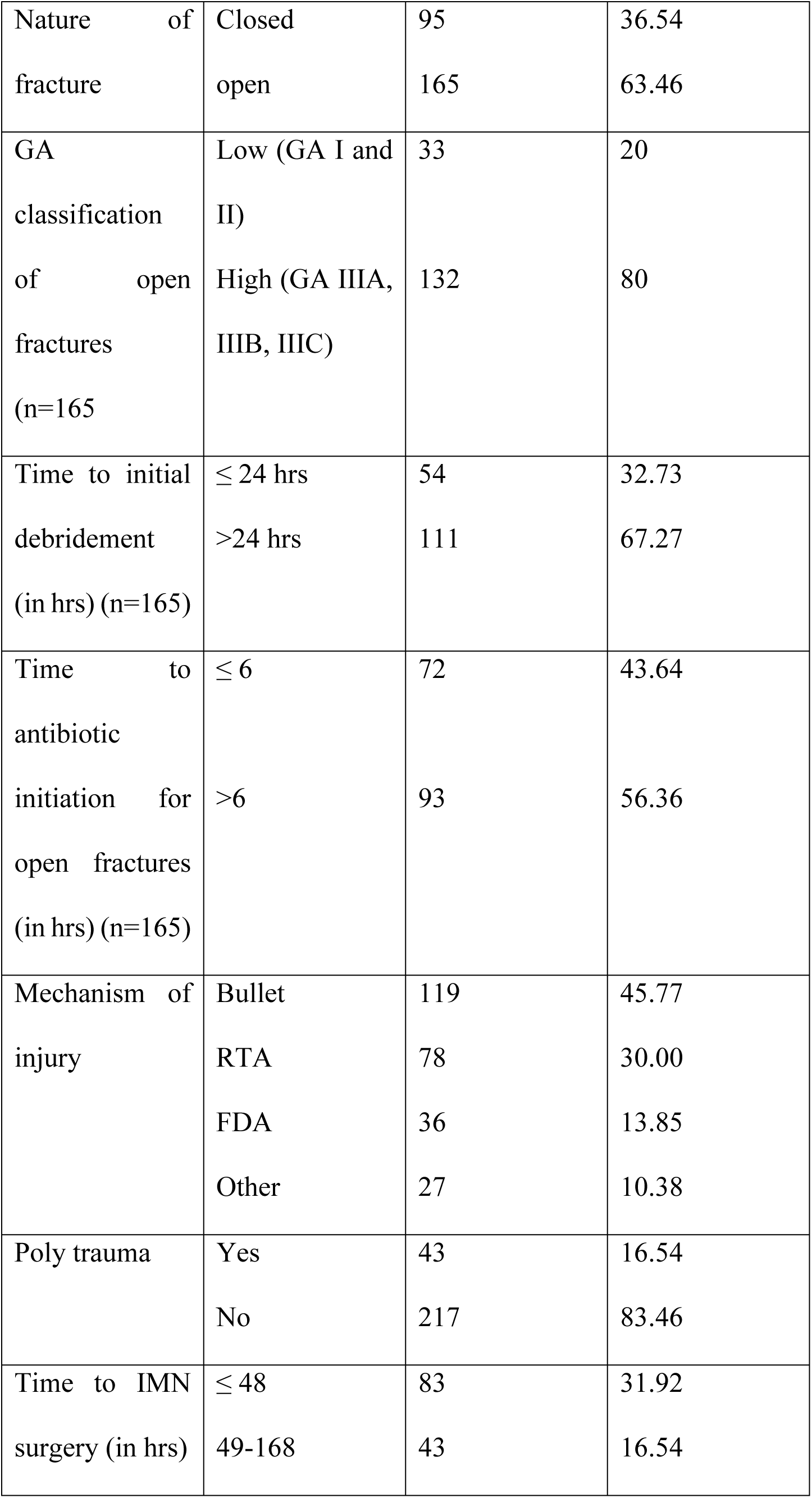
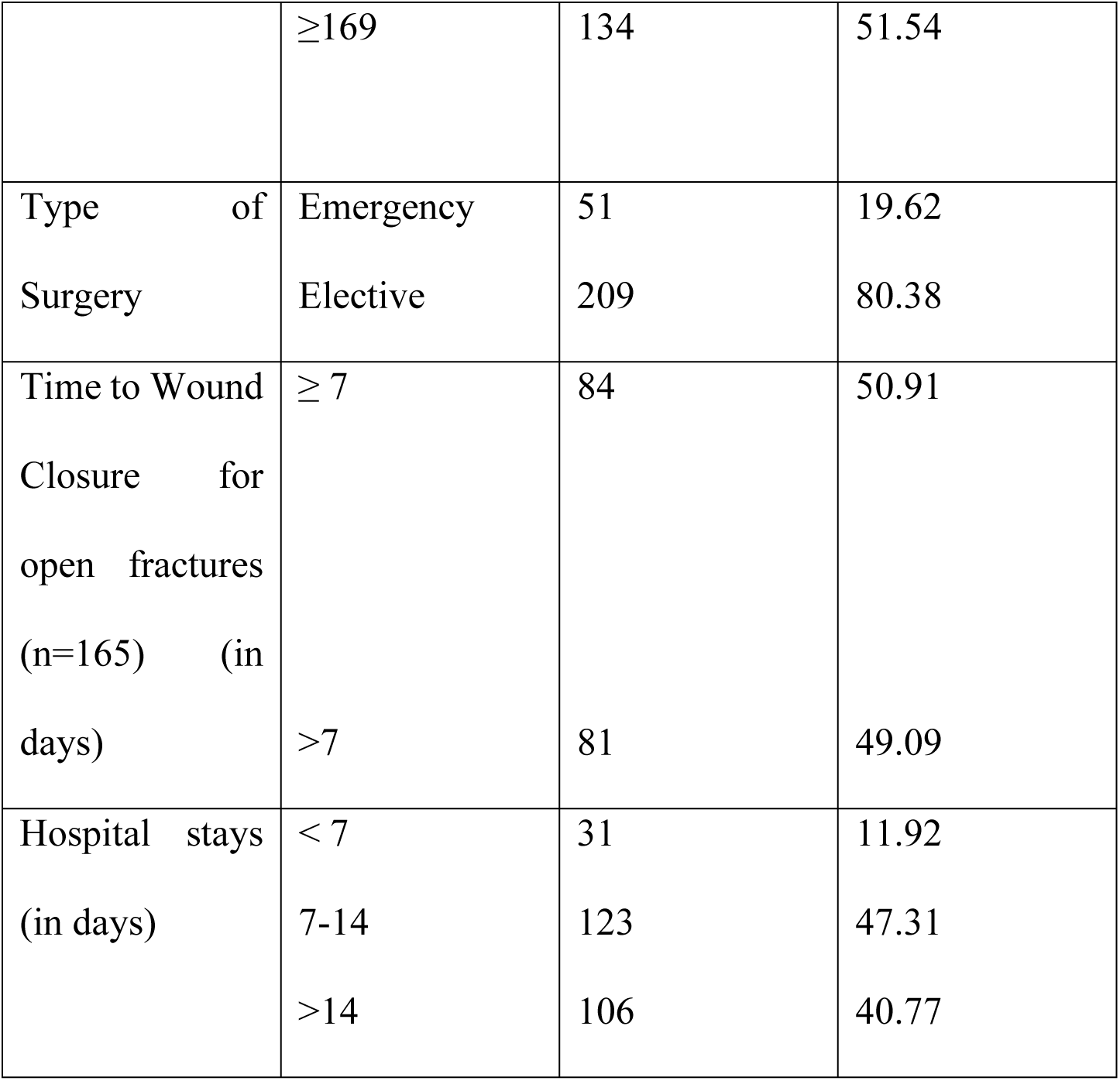
Demographic and Clinical characteristics of study participants.

| Variable | Category | Frequency<br>(n=260) | Percent (%) |
| --- | --- | --- | --- |
| Age group<br>(years) | 18-30 | 153 | 58.85 |
|  | 31-60 | 90 | 34.62 |
|  | >61 | 17 | 6.54 |
| Sex | Male | 217 | 83.46 |
|  | Female | 43 | 16.54 |
| Residence | Rural | 196 | 75.38 |
|  | Urban | 64 | 24.62 |
| Occupation | Farmer | 124 | 47.69 |
|  | Merchant | 21 | 8.08 |
|  | Student | 43 | 16.54 |
|  | Housewife | 38 | 14.62 |
|  | Other | 34 | 13.08 |
| Duration of<br>injury at<br>presentation (in<br>hrs) | $\geq 12$ | 131 | 50.38 |
|  | 13-48 | 92 | 35.38 |
| | $\geq 49$ | 37 | 14.23 |
| Initial<br>Hemoglobin<br>level (g/dl) | $\leq 10$ | 198 | 76.15 |
| | $< 10$ | 62 | 23.85 |
| Bone fractured | Tibia | 95 | 36.54 |
|  | Femur | 165 | 63.46 |
| Nature of fracture | Closed | 95 | 36.54 |
|  | open | 165 | 63.46 |
| GA classification of open fractures (n=165) | Low (GA I and II) | 33 | 20 |
|  | High (GA IIIA, IIIB, IIIC) | 132 | 80 |
| Time to initial debridement (in hrs) (n=165) | $\leq 24$ hrs | 54 | 32.73 |
| | $>24$ hrs | 111 | 67.27 |
| Time to antibiotic initiation for open fractures (in hrs) (n=165) | $\leq 6$ | 72 | 43.64 |
| | $>6$ | 93 | 56.36 |
| Mechanism of injury | Bullet | 119 | 45.77 |
|  | RTA | 78 | 30.00 |
|  | FDA | 36 | 13.85 |
|  | Other | 27 | 10.38 |
| Poly trauma | Yes | 43 | 16.54 |
|  | No | 217 | 83.46 |
| Time to IMN surgery (in hrs) | $\leq 48$ | 83 | 31.92 |
|  | 49-168 | 43 | 16.54 |
| | $\geq 169$ | 134 | 51.54 |
| Type of Surgery | Emergency | 51 | 19.62 |
|  | Elective | 209 | 80.38 |
| Time to Wound Closure for open fractures (n=165) (in days) | $\geq 7$ | 84 | 50.91 |
| | $> 7$ | 81 | 49.09 |
| Hospital stays (in days) | $< 7$ | 31 | 11.92 |
|  | 7-14 | 123 | 47.31 |
| | $> 14$ | 106 | 40.77 |

Clinical Characteristics of the study were organized thematically to enhance clarity and relevance. The domains included duration of injury presentation, hemodynamic status, injury profile, perioperative management, surgical timing, post operative wound care, and hospital course. Variables were summarized using frequencies and percentages, with categorization applied to skewed distributions to support both analytic precision and clinical interpretation (Table 1). The median age of presentation was 12 hrs (IQR: 5.5 hrs – 24 hrs). The distribution was highly right-skewed (skewness = 5.53). (Figure 2). Categorizing duration of injury presentation into three (within 12 hrs, 13-48 hrs and after 48 hrs), 131(50.38 %) of cases arrived at UOG CSH within 12 hrs of injury whereas 37 cases (14.23 %) came after two days of injury. The hemoglobin level at presentation ranged from 6.0 to 15.8 g/dL, with a mean of 11.57 ± 2.34 g/dL and a median of 11.8 g/dL. Among the 260 participants, 64 patients (24.6%) had hemoglobin levels below 10 g/dL, suggesting a notable burden of anemia at the time of admission, which may influence infection risk and wound healing outcomes (Figure 3).

**Figure 2:**
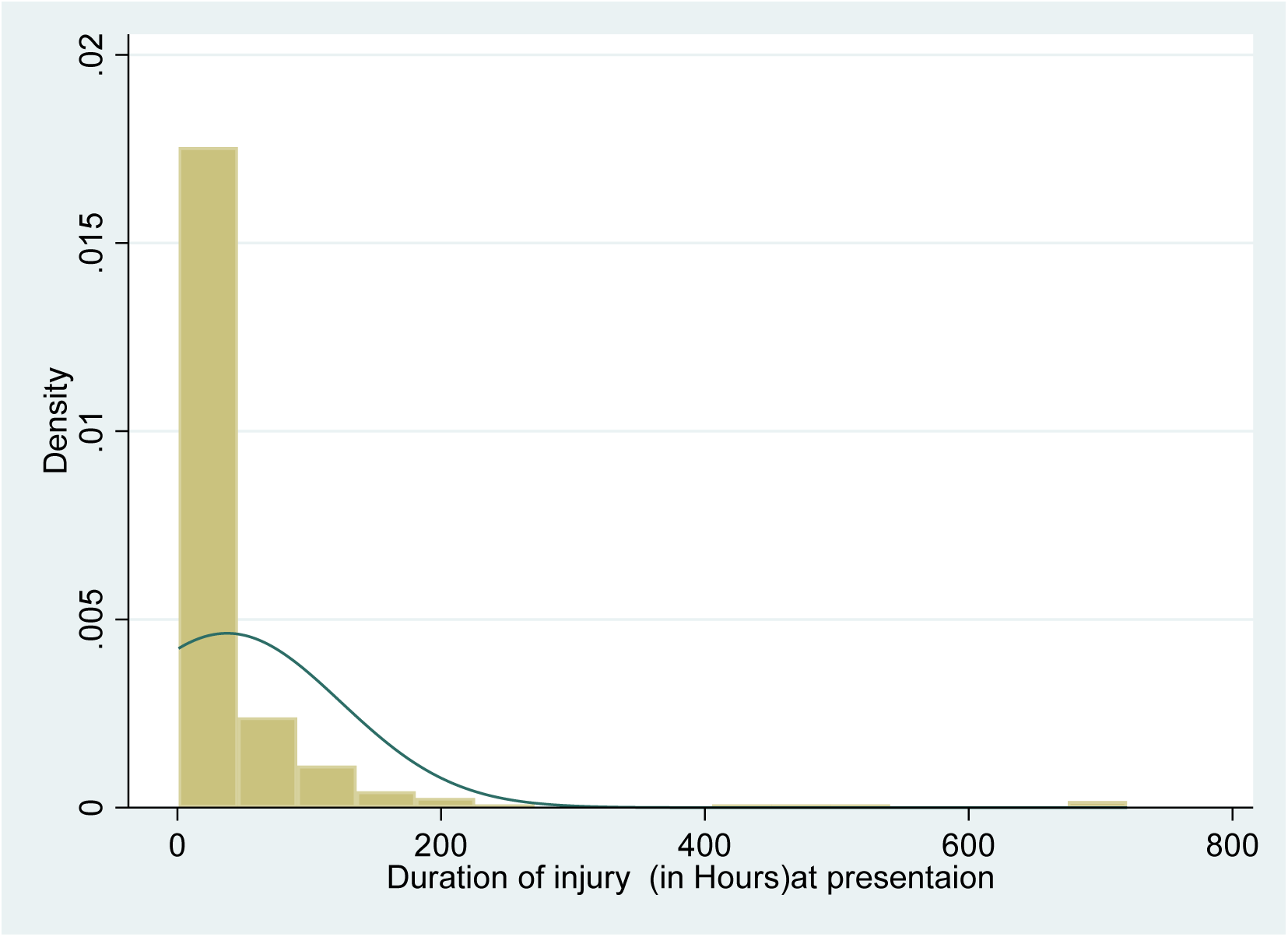
Histogram showed distribution of duration of presentation following injury.

**Figure 3:**
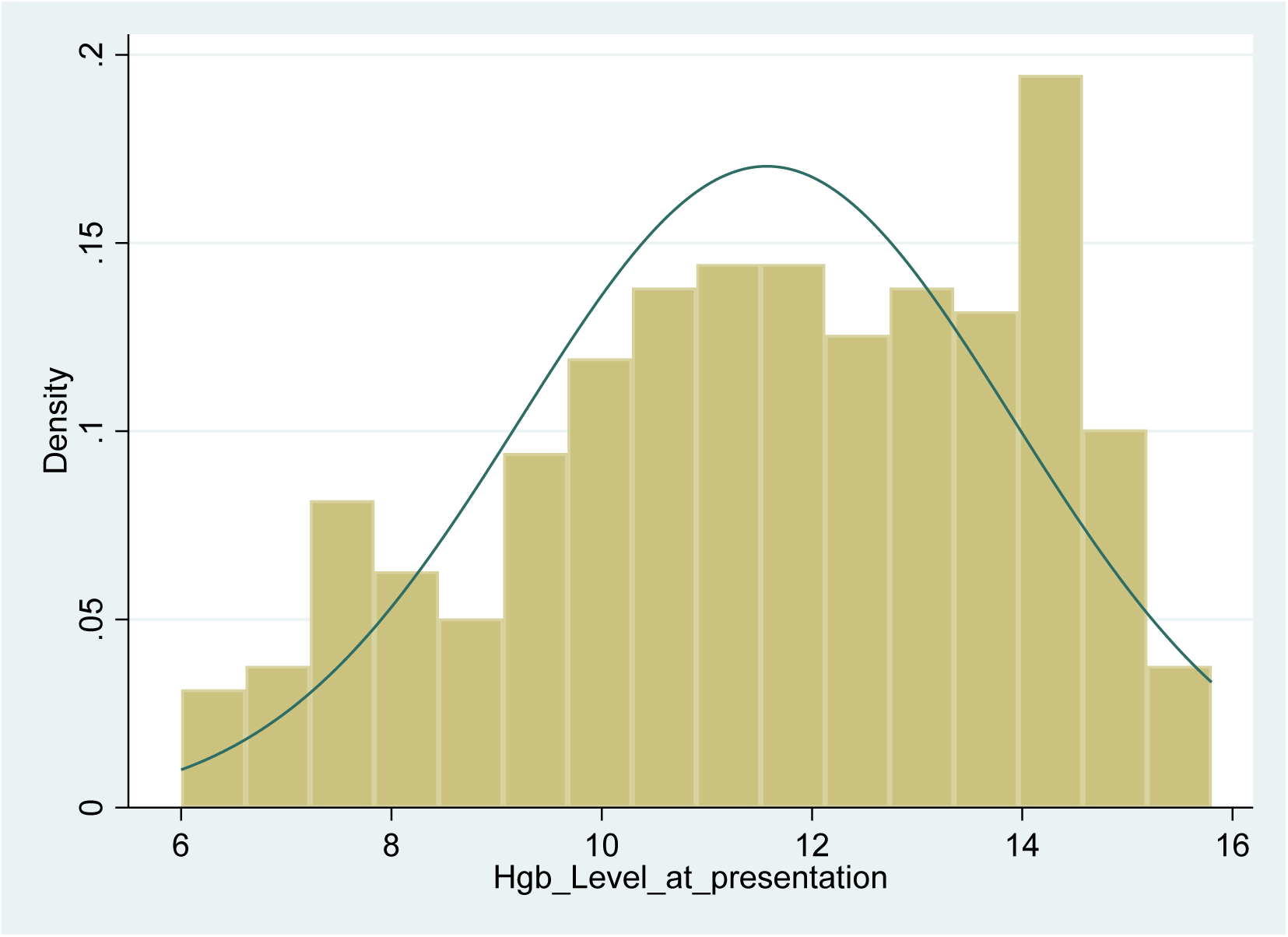
Initial Hemoglobin level (g/dl) at presentation.

Of the 260 patients who underwent IMN procedures in the tibia or femur, 165 (63.46%) had femoral fractures, while the remaining 95 (36.54%) sustained tibial fractures. A total of 165 cases (63.46%) were classified as open fractures. Among these, 132 patients (80%) presented with high-grade open fractures, corresponding to Gustilo-Anderson classifications IIIA, IIIB, or IIIC (Table 1).

Of the 165 patients with open fractures, 72 (43.64%) received prophylactic antibiotics within 6 hours of injury, while the remaining 93 (56.36%) received antibiotics after 6 hours. In contrast, a large majority—111 patients (67.27%)—underwent initial surgical debridement more than 24 hours after injury, indicating substantial delays in early wound management (Table 1). Of the 260 patients who underwent IMN surgery for tibial or femoral fractures, bullet injuries accounted for the largest proportion, affecting 119 individuals (45.77%). Road traffic accidents (RTAs) were the second most common mechanism, responsible for 78 cases (30.00%). Polytrauma was documented in 43 patients (16.54%), while the remaining cases involved isolated fractures of either the tibia or femur. These distributions are summarized in Table 1 and visually depicted in Figure 4.

**Figure 4:**
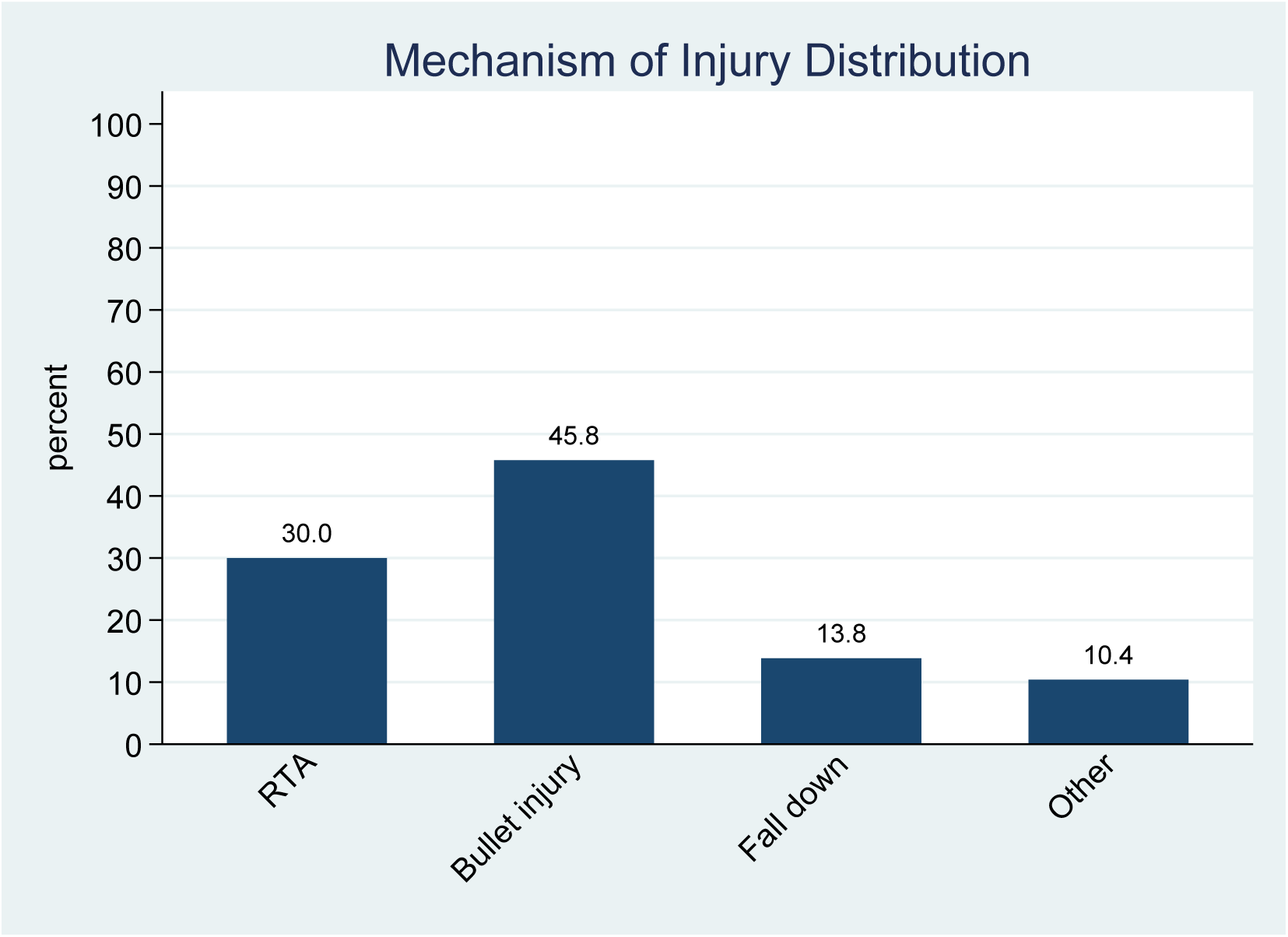
Percentage distribution of mechanism of injury among IMN patients.

The time to IMN surgery ranged from 6 to 1008 hours, with a median of 180 hours and a mean of 212.6 ± 177.9 hours. The distribution was positively skewed, indicating that many patients experienced delays beyond the median (Figure 5). For clinical interpretability, surgical timing was also categorized into early (≤48 hours), delayed (49–168 hours), and late (≥169 hours) groups(20). Over half of the patients (51.54%) underwent late surgery, while only 31.92% received early intervention (Table1). In addition, 209 (80.38 %) out of 260 IMN surgeries were elective and the remaining 51 (19.62 %) were operated on an emergency basis (Table 1). Among the 165 patients with open fractures, 84 (50.91%) underwent wound closure within 7 days of injury, while the remaining 81 (49.09%) had their wounds closed after 7 days (Table 1).

**Figure 5:**
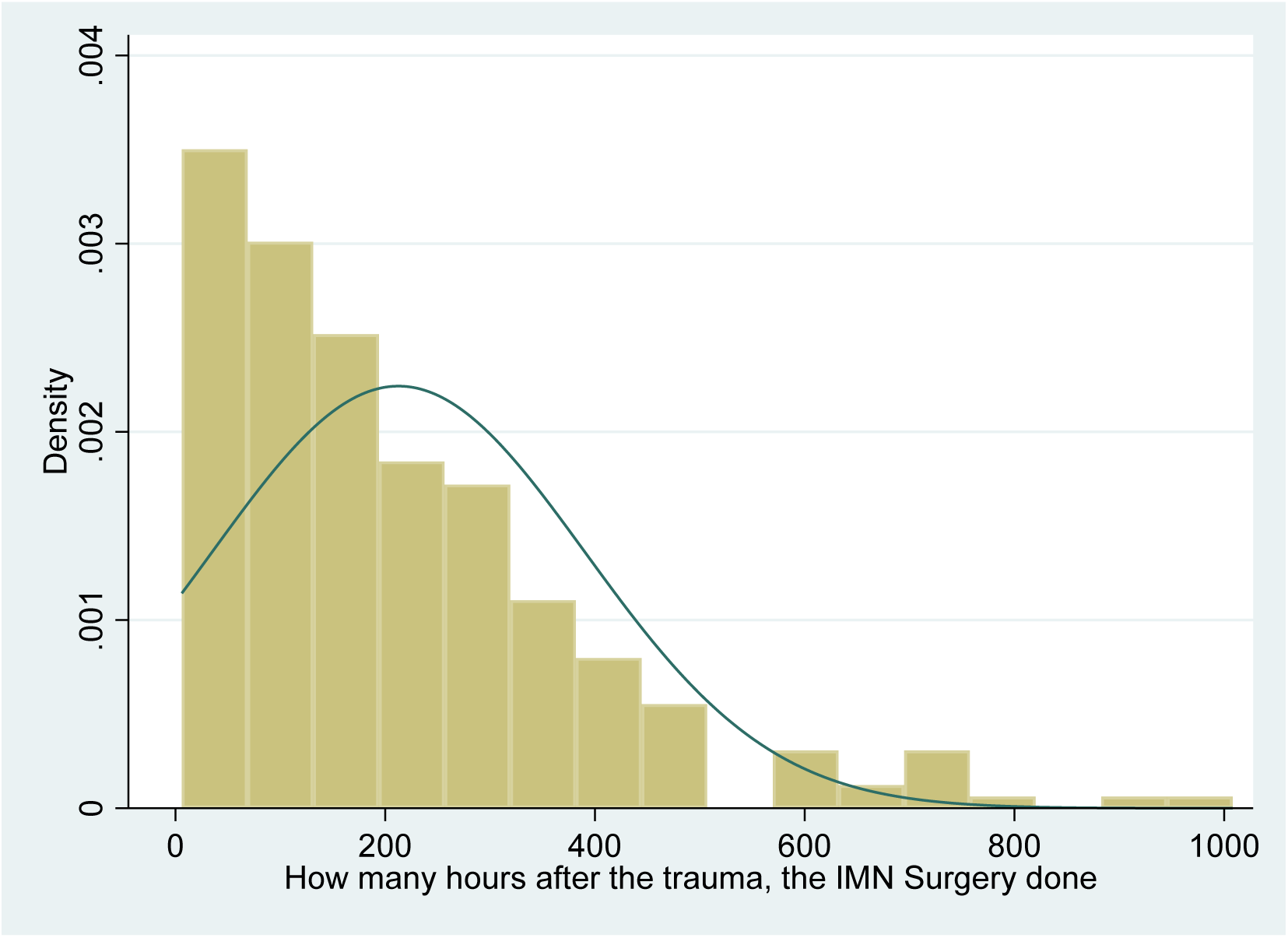
Timing of definitive IMN surgery in hours from trauma.

The duration of hospital stays among IMN-treated patients ranged from 3 to 60 days, with a median of 13 days and a mean of 14.85 days (SD ±8.74). The interquartile range (IQR) was 9 to 19 days, indicating moderate variability (Figure 6). Notably, 11.92% of patients were discharged within 7 days, while 47.31% stayed between 7 and 14 days, and 40.77% remained hospitalized for more than 14 days (Table 1).

**Figure 6:**
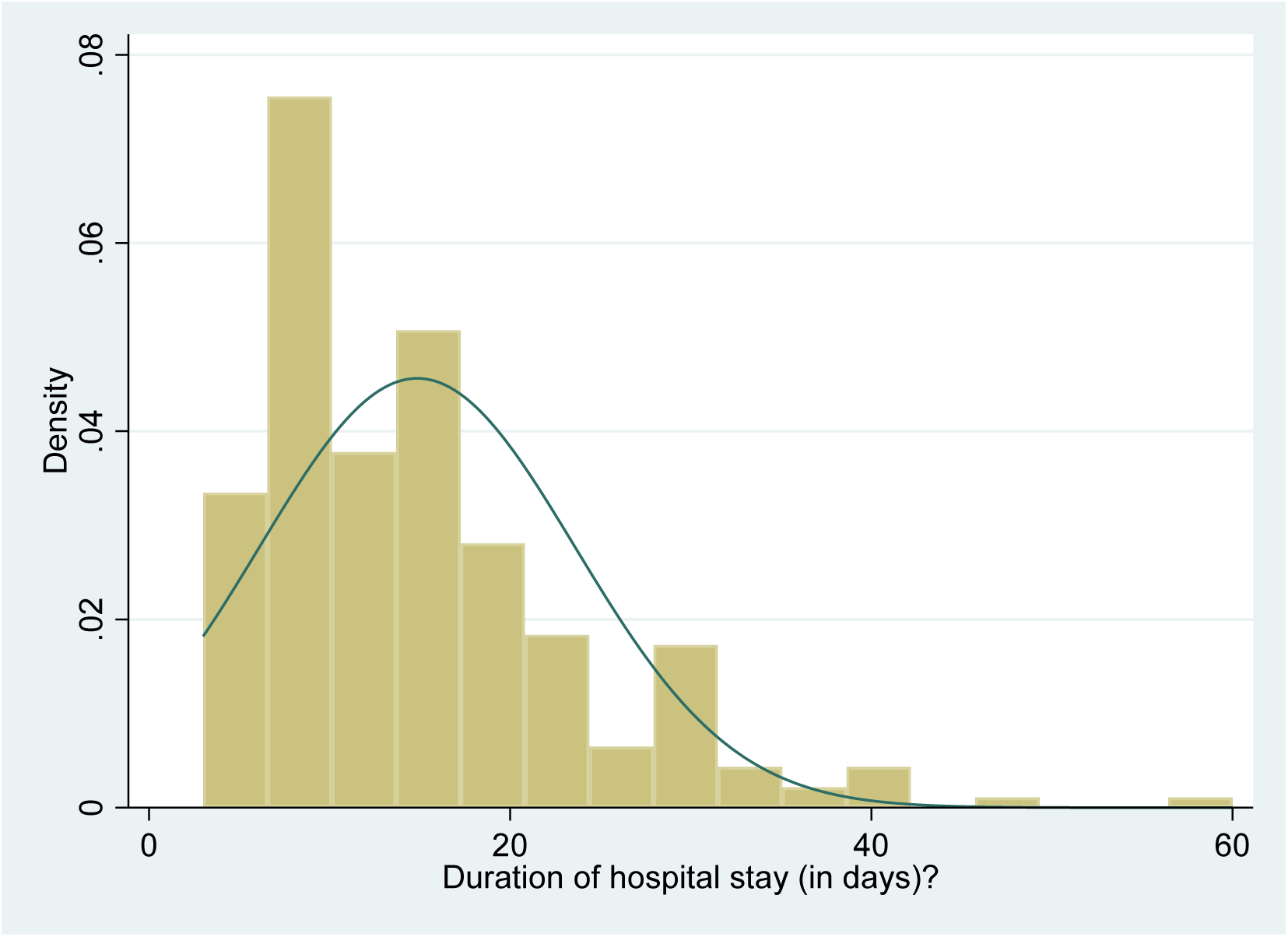
Duration of hospital stay in days.

Bullet injury was the predominant cause, followed by road traffic accidents, falls, and other trauma. However, bullet injuries were excluded from the regression analysis due to their low frequency and distinct characteristics.

### Magnitude of Implant-associated Infections Following IMN Surgery

Among 260 patients who underwent IMN surgery for tibial or femoral fractures, 22 cases of implant-associated infection were documented, yielding an overall infection rate of 8.46% (95% CI: 5.38%– 12.53%) (Figure 7). Of these infections, 12 cases out of 260 (4.62%) were classified as deep infections, while the remaining 10 out of 260 (3.85 %) were superficial.

**Figure 7:**
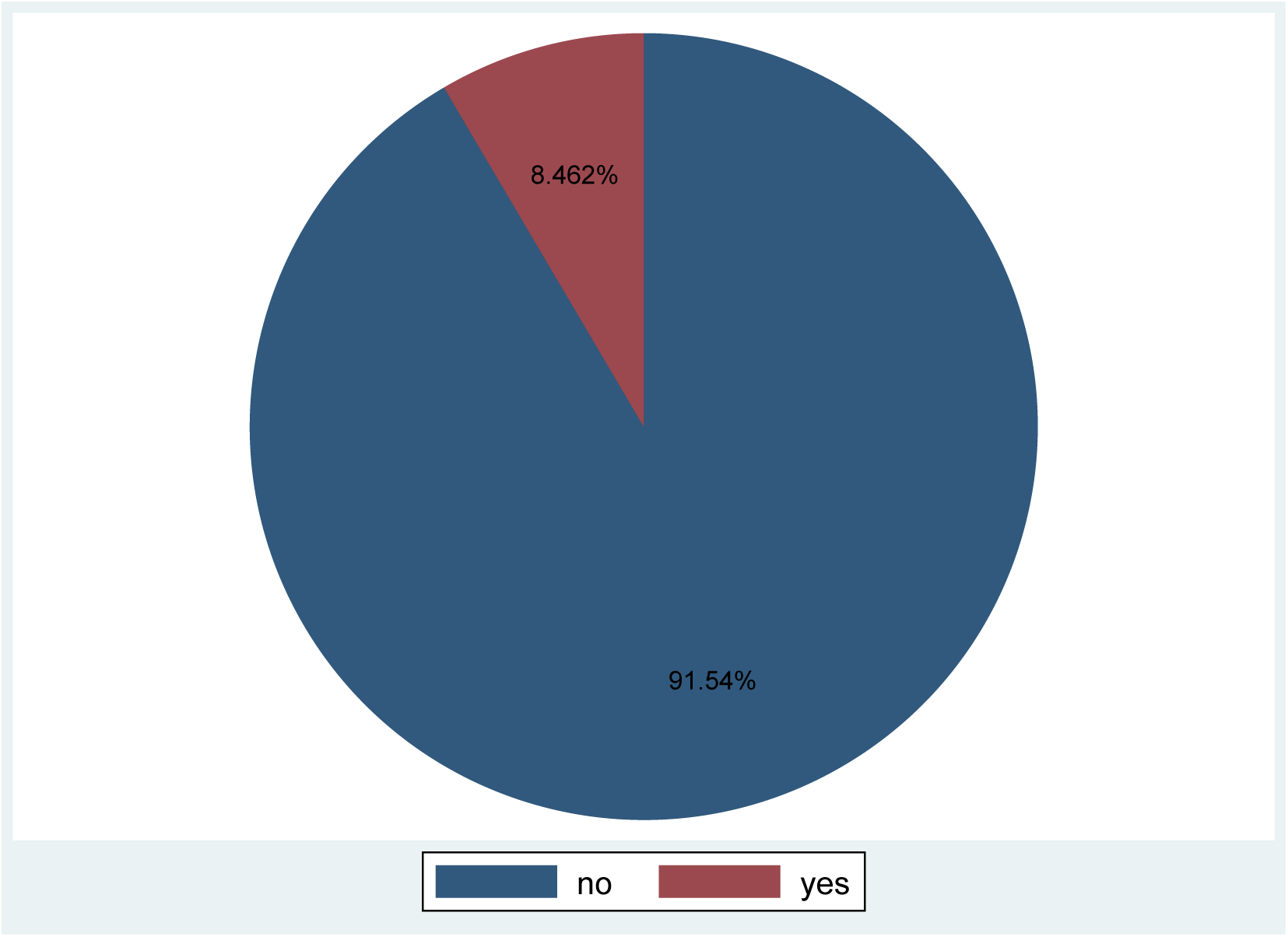
Proportion of IMN-associated infections.

### Determinants of IMN-associated Infections

The bivariate analysis identified several variables with potential associations to IMN-associated infections, using a liberal threshold of p ≤ 0.2 to capture clinically relevant trends. These variables included age category, duration of injury at presentation, nature of fracture, Gustilo-Anderson classification of open fractures, time to initial debridement, mechanism of injury, polytrauma status, type of IMN surgery, and duration of hospital stay (Table 2).

**Table 2:** Bivariate Analysis of Determinants of IMN-associated Infection.

| Variable | Category | Infection status |  | Crude OR<br>(95 % CI) | P-value |
| --- | --- | --- | --- | --- | --- |
|  |  | Yes<br>n | No<br>n |  |  |
| Sex | Male | 18 | 199 | 0.09 (0.056-0.146) | 0.829 |
|  | Female | 4 | 39 | 0.10 (0.037-0.287) |  |
| Age (years) | 18-30 | 17 | 136 | 0.125(0.076-0.206) | **0.048** |
|  | 31-60 | 5 | 85 | 0.059(0.024-0.144) |  |
|  | ≥ 61 | 0 | 17 |  |  |
| Residence | Rural | 17 | 179 | 0.095(0.058-0.156) | 0.830 |
|  | Urban | 5 | 59 | 0.085(0.034-0.211) |  |
| Occupation | Farmer | 10 | 114 | 0.088 (0.046-0.167) | 0.620 |
|  | Merchant | 1 | 20 | 0.050 (0.006-0.373) |  |
|  | Student | 3 | 40 | 0.075(0.023-0.242) |  |
|  | Housewife | 3 | 29 | 0.103(0.031-0.340) |  |
|  | Other | 4 | 33 | 0.121(0.043-0.342) |  |
| Duration of injury at presentation (in hrs) | ≤12 | 17 | 114 | 0.149(0.089-0.248) | **0.005** |
|  | 13-48 | 5 | 87 | 0.057(0.023-0.142) |  |
|  | ≥49 | 0 | 37 | 0.000 |  |
| Initial hemoglobin level (g/dl) | ≥10 | 15 | 183 | 0.082(0.048-0.139) | 0.360 |
|  | < 10 | 7 | 55 | 0.127(0.058-0.279) |  |
| Type of bone fractured | Femur | 15 | 150 | 0.100(0.059-0.170) | 0.631 |
|  | Tibia | 7 | 88 | 0.080(0.037-0.172) |  |
| Nature of the fracture | Open | 20 | 145 | 0.138(0.086-0.220) | **0.005** |
|  | Closed | 2 | 93 | 0.021(0.005-0.087) |  |
| GA Classification | Lower class (Type I and Type II) | 1 | 32 | 0.31 (0.004-0.229) | 0.075 |
|  | Higher class (GA IIIA, B, C) | 19 |  | 0.168(0.103-0.273) |  |
| Time to initial debridement (in hrs) | ≤24 | 11 | 43 | 0.256(0.131-0.496) | **0.024** |
|  | >24 | 9 | 102 | 0.088(0.044-0.174) |  |
| Time to antibiotic initiation for | ≤6 | 11 | 61 | 0.180(0.095-0.343) | 0.276 |
|  | >6 | 9 | 84 | 0.107(0.054-0.213) |  |
| open fractures<br>(in hrs) |  |  |  |  |  |
| Mechanism of injury | RTA | 5 | 73 | 0.068(0.028-0.169) | 0.147 |
|  | Bullet injury | 17 | 102 | 0.167(0.100-0.279) |  |
|  | Fall down injury | 0 | 36 | 0.000 |  |
|  | other | 0 | 27 | 0.000 |  |
| polytrauma | Yes | 1 | 42 | 0.107(0.068-0.168) | 0.1 |
|  | No | 21 | 196 | 0.024(0.003-0.173) |  |
| Type of IMN surgery | Emergency | 9 | 42 | 0.214(0.104-0.440) | **0.008** |
|  | Elective | 13 | 196 | 0.066(0.038-0.116) |  |
| Time to IMN surgery (in hrs) | ≤48 | 14 | 69 | 0.202 (0.114-0.360) | **0.002** |
|  | 49-168 | 2 | 41 | 0.049(0.012-0.202) |  |
|  | >168 | 6 | 128 | 0.047(0.021-0.106) |  |
| Time to wound closure for open fractures (in days) | ≤7 | 11 | 73 | 0.150 (0.080-0.284) | 0.697 |
|  | >7 | 9 | 72 | 0.125(0.063-0.250) |  |
| Duration of Hospital stays (in days) | <7 | 1 | 30 | 0.33(0.005-0.244) | 0.059 |
|  | 7-14 | 8 | 115 | 0.100(0.034-0.142) |  |
|  | >14 | 13 | 93 | 0.140(0.078-0.250) |  |

**Table 3:**
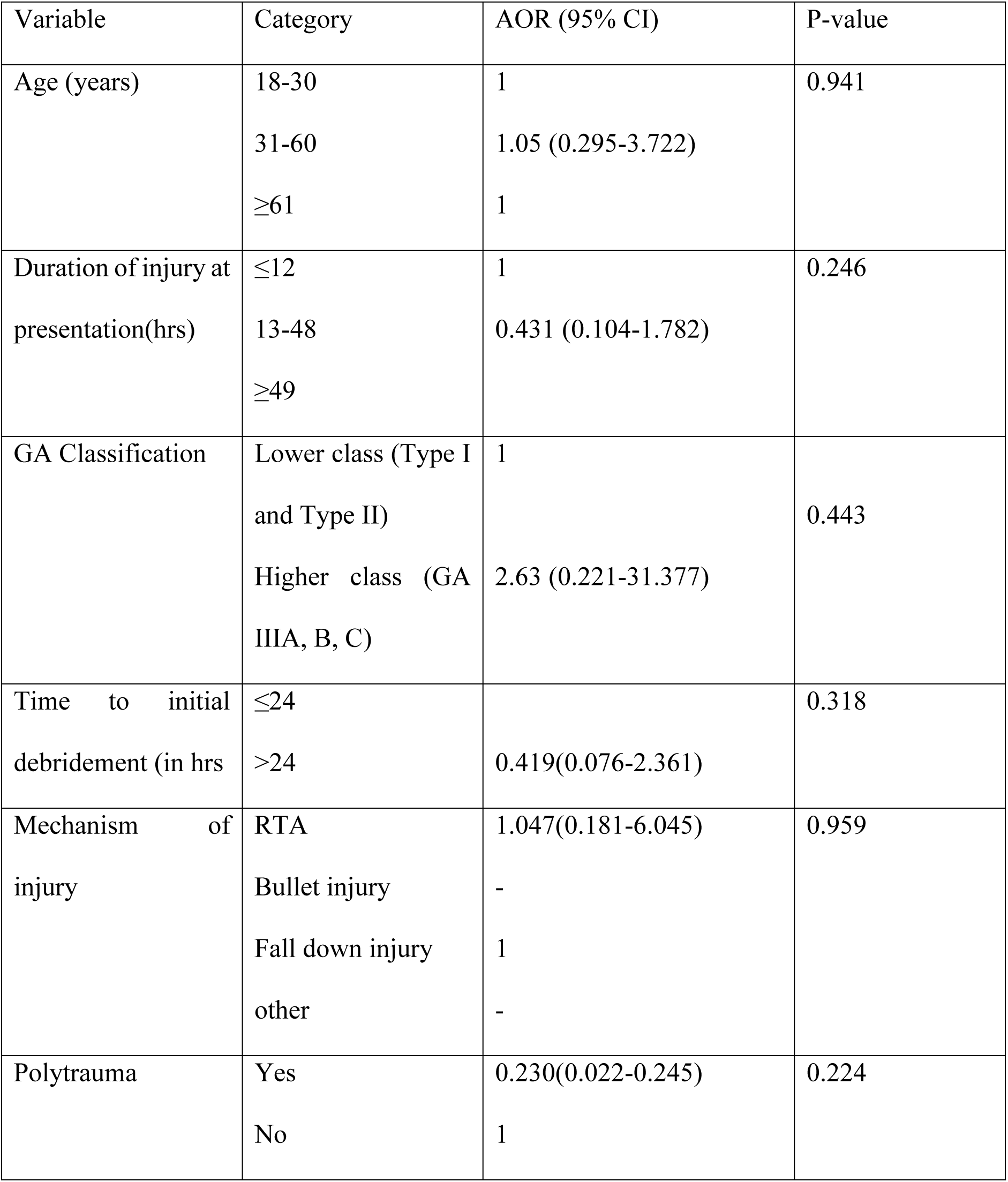
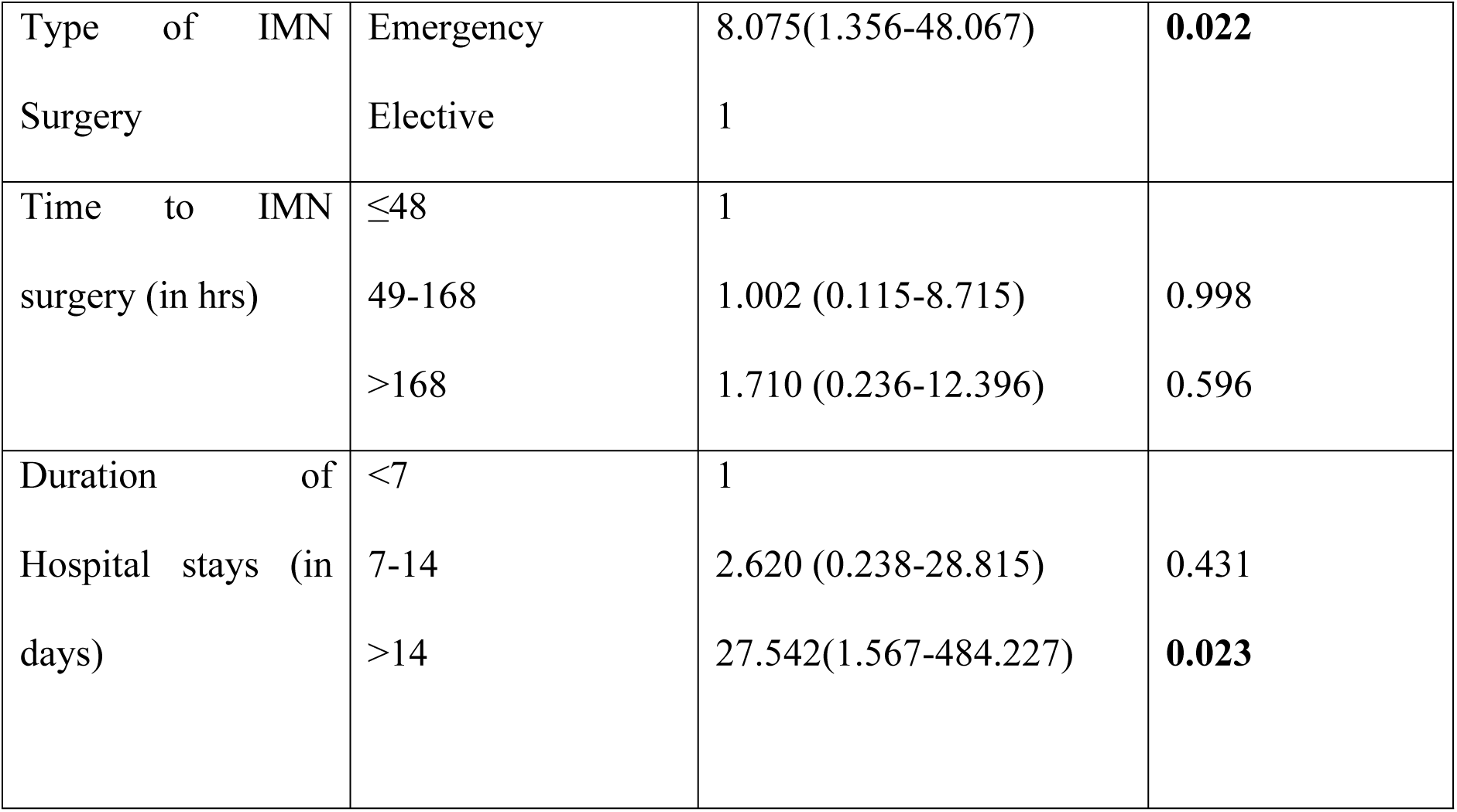
Multivariate Logistic Regression Analysis of Determinants Associated with Implant-associated Infections Following IMN.

In the multivariate logistic regression model, two factors demonstrated statistically significant associations with IMN-associated infection. Patients who underwent emergency IMN surgery had eight times higher odds of developing infection compared to those treated electively (AOR = 8.075; 95% CI: 1.356–48.067; p = 0.022). Similarly, patients who remained hospitalized for more than 14 days had 27 times higher odds of infection compared to those discharged within 7 days (AOR = 27.542; 95% CI: 1.567– 484.227; p = 0.023) (Table 3).

Out of 22 infected cases, culture and sensitivity testing was performed in 9 (40.9%). Of these, 4 (44.4%) yielded growth. The most common pathogen was E. coli (50%), followed by K. pneumoniae (25%) and non-lactose fermenting Gram-negative rods (25%). The isolates showed sensitivity to meropenem, ciprofloxacin, chloramphenicol, gentamycin, imipenem, and ceftazidime, while resistance was observed against ceftriaxone, ampicillin, azithromycin, vancomycin, cefuroxime, cotrimoxazole, ciprofloxacin, and piperacillin-tazobactam (Table 4).

**Table 4:**
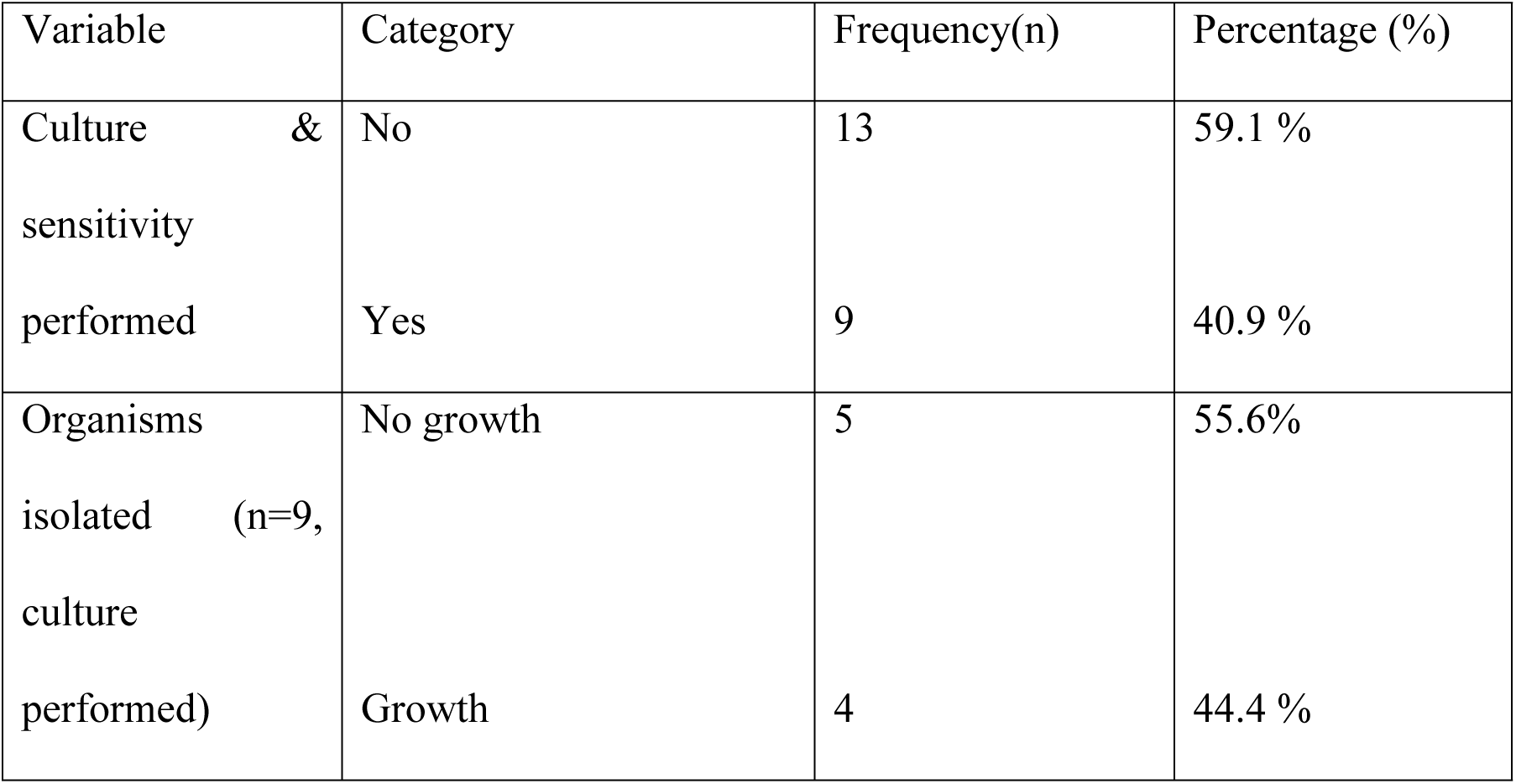
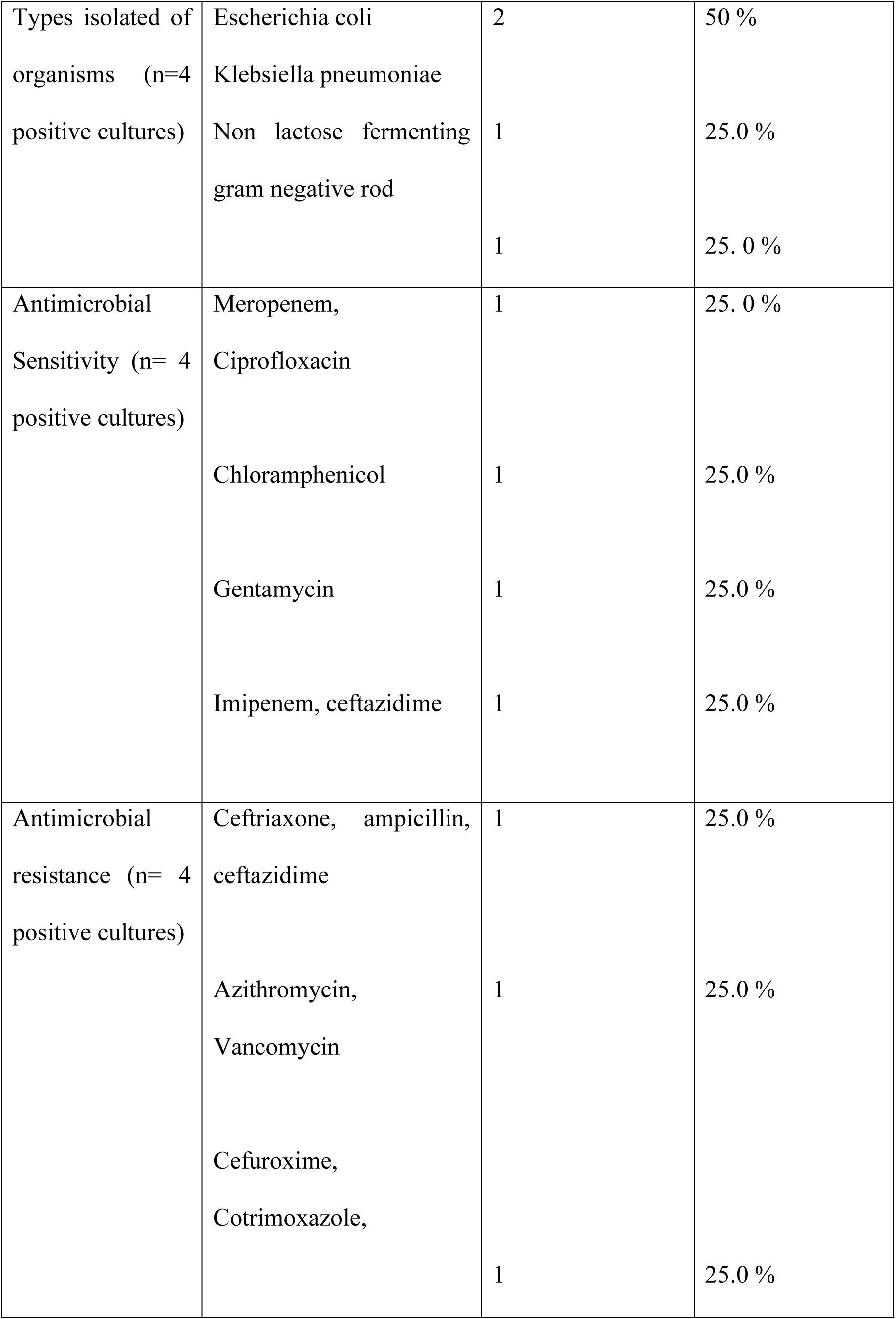
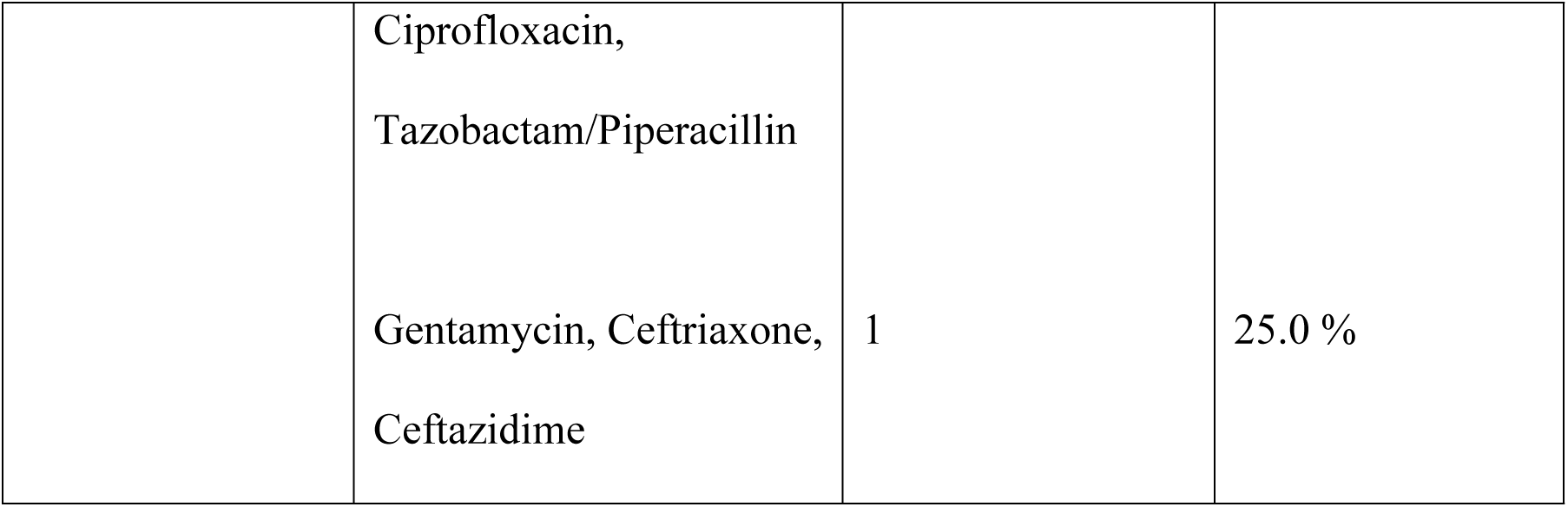
Microbiological Profile and Antimicrobial Resistance Patterns of Pathogens Isolated from IMN-Associated Infections (n = 22)

## Discussion

This study found an overall implant-associated infection rate of 8.46 % (95 % CI: 5.38 %– 12.53 %) following IMN surgery in the tibia or femur, with deep infections constituting 4.62 % (n = 12) and superficial infections 3.85 % (n = 10).

Compared with Beza et al (17), who reported a 4.18 % infection rate after long-bone IMN, our observed rate is substantially higher. This discrepancy likely reflects the greater prevalence of open fractures in our cohort (approximately 64 % [165/260] versus 24 % in Beza et al.

A retrospective cross-sectional study at Saint Paul’s Hospital Millennium Medical College in Addis Ababa similarly reported an overall infection rate of 9.3 % (18), which aligns closely with our findings. However, the prevalence of deep infections in our series (4.62 %) exceeded that reported by Desta et al. (3.45 %) (Desta et al. 2023). This difference may stem from fracture severity: GA III A injuries predominated in our study, whereas GA II fractures were most common in Saint Paul’s cohort.

By contrast, Young et al(7) documented a markedly lower overall IMN infection rate of 1 %. Their study’s reliance on an incomplete follow-up registry (Sign Online Surgical Database) may have underestimated true infection rates. Similarly low rates have been observed in high-resource settings(21–23), reflecting differences in perioperative environments, operating room sterility protocols, and patient baseline characteristics. Moreover, those studies did not focus on specific bone or implant types, limiting direct comparability.

Conversely, several reports describe higher infection rates after IMN surgery to long bones of lower extremity than ours—11.9 % in Whiting et al. and 12.9 % in Adesina et al(24,25). These elevated rates are attributable to study populations confined to open tibial shaft fractures (25) or combined open femoral and tibial shaft fractures(24), both of which carry greater contamination risk and soft-tissue compromise.

In the present study, multivariate analysis identified emergency surgery and prolonged hospital stay exceeding two weeks as significant predictors of infection following IMN surgery of femoral and tibial fractures (Table 3). The association between prolonged hospitalization and increased risk of infection is consistent with the findings of Weaver et al(26), who reported that a hospital stay of more than 10 days was statistically significant in predicting orthopedic surgical site infections. Similarly, a prospective study of the bacteriological profile and risk factors of infection after internal fixation of closed fractures of long bones identified hospitalization for more than two weeks is an independent risk factor for the development of infection(27). Extended hospitalization may predispose patients to infection through prolonged exposure to nosocomial pathogens, repeated invasive procedures, and overall increased vulnerability due to comorbid conditions.

Similarly, our finding that emergency surgery is an independent risk factor for infection aligns with the results of Rezae et al (28), who demonstrated that emergency IMN for femoral and tibial shaft fractures was associated with a higher risk of postoperative infection. This may be due to higher wound contamination risk and limited preoperative preparation in emergency surgeries.

However, the role of emergency surgery as a determinant of infection remains controversial. In contrast to our findings, studies conducted by Ercole et al. and Maksimovic et al. did not identify emergency surgery as an independent predictor of infection in orthopedic operations(29,30). These discrepancies may reflect differences in study populations, case selection, perioperative protocols, and infection control measures across institutions

In our study, the number of positive bacterial cultures was relatively small. Among these, all isolates were gram-negative organisms, predominantly Escherichia coli, Klebsiella pneumoniae, and non-lactose-fermenting gram-negative rods (Table 4). This finding contrasts with the results of other studies, where Staphylococcus species were reported as the most frequently isolated pathogens, followed by gram-negative organisms(27,30,31). Several factors may explain this discrepancy. First, in many cases, cultures were obtained after the initiation of antibiotic therapy, which may have suppressed the growth of gram-positive bacteria. Second, the limited availability of culture media in our setting may have restricted the growth of a wider range of organisms. Finally, contamination during the collection or processing of specimens could have influenced the spectrum of organisms identified. These findings highlight the need for timely culture sampling, improved microbiological facilities, and strict adherence to aseptic techniques, as they are essential for guiding appropriate antibiotic selection and improving infection management in our hospital.

### Limitations

This study has several limitations that should be acknowledged. First, its retrospective cross-sectional design relied on secondary data from medical records, which may have been incomplete or inconsistently documented. This could have led to missing information on relevant variables such as nutritional status, glycemic control, and smoking history, which are known to influence infection risk. Second, culture and sensitivity testing were performed in less than half of the infected cases, limiting the ability to fully characterize the microbiological spectrum and antimicrobial resistance patterns. Third, lack of standardized follow-up likely led to underestimation, highlighting the need for prospective studies. Finally, as the study was conducted in a single tertiary hospital, the findings may not be generalizable to other institutions with different patient populations, perioperative protocols, or resource settings.

## Conclusion

This study demonstrated that the overall infection rate following intramedullary nailing of femoral and tibial fractures at UOG CSH was 8.46%, which is higher than rates reported in some high-resource settings but comparable to findings from other Ethiopian and regional studies. Multivariate analysis identified emergency surgery and prolonged hospital stay (>14 days) as independent predictors of infection. Furthermore, although the culture yield was low, all positive isolates were gram-negative organisms, predominantly Escherichia coli and Klebsiella pneumoniae, in contrast to the staphylococcal predominance reported in other studies. These findings underscore the need for timely culture sampling, optimization of perioperative protocols, and strategies to minimize unnecessary hospital stays to reduce infection risk.

### Future Directions

Future studies should adopt prospective cohort or longitudinal designs to allow more robust assessment of causality and better capture of late-onset infections. Expanding microbiological capacity, including the use of diverse culture media and molecular diagnostic tools, would help to more accurately define the spectrum of causative pathogens and their resistance patterns. Multicenter studies across Ethiopia and similar low-resource settings are also warranted to enhance generalizability and identify institution-specific infection control gaps. Additionally, intervention-focused research aimed at reducing delays to surgery, improving early wound management, and strengthening antibiotic stewardship will be critical for lowering the burden of implant-associated infections in this context.

## Declarations

### Ethics approval and Consent to participate

Ethical approval was obtained from the Institutional Review Board (IRB) of the University of Gondar. Administrative permissions were also secured from the hospital management. The study was conducted in accordance with the Declaration of Helsinki.

### Consent for publication

Not applicable, as this study did not include identifiable individual patient data.

### Availability of data and materials

The datasets used and/or analyzed during the current study are available from the corresponding author on reasonable request.

### Authors’ contributions

AMZ conceived and designed the study, conducted the data collection, performed the data analysis and interpretation, and drafted the initial manuscript.

EBK contributed to the study design, assisted in data acquisition, and critically revised the manuscript.

BTA contributed to data analysis, interpretation of results, and substantive manuscript revision.

SSZ assisted with data collection, contributed to data interpretation, and manuscript revision.

KTA provided clinical input, contributed to interpretation of findings, and critically revised the manuscript.

YDM contributed to the study design, provided surgical expertise, and revised the manuscript for important intellectual content.

All authors read and approved the final version of the manuscript and agree to be accountable for all aspects of the work.

## Data Availability

All relevant data are within the manuscript and its Supporting Information files. Patient-level data have been de-identified to protect confidentiality in accordance with ethical approval.

## List of abbreviations

AOR: Adjusted Odds Ratio
CDC: Center for disease control and Prevention
CI: Confidence interval
CSH: Comprehensive Specialized Hospital
FDA: Fall Down Accident
g/dl: gram per deciliter
GA: Gustilo –Anderson
Hrs: hours
IMN: Intramedullary Nailing
IQR: Interquartile Range
IRB: Institutional Review Board
LMICs: Low– and middle-income countries
N: number
OR: odds Ration
RTA: Road Traffic Accident
SD: Standard Deviation
UOG: University of Gondar
VIF: Variance inflation factor

## Acknowledgements

Not applicable

